# Transfer Learning for Neuroimaging via Re-use of Deep Neural Network Features

**DOI:** 10.1101/2022.12.11.22283324

**Authors:** Peter Holderrieth, Stephen Smith, Han Peng

## Abstract

A major problem in the application of machine learning to neuroimaging is the technological variability of MRI scanners and differences in the subject populations across studies. Transfer learning (TL) attempts to alleviate this problem. TL refers to a family of methods which acquire knowledge from related tasks to improve generalization in the tasks of interest. In this work, we pre-train a deep neural network on UK Biobank MRI data on age and sex prediction, and study the predictions of the network on three small MRI data sets. We find that the neural networks can extract meaningful features from unseen data sets under the necessary but also sufficient condition that the network was pre-trained to predict the label of interest (e.g. pre-trained on age prediction if age prediction is the task of interest). Based on this, we propose a transfer learning pipeline which relies on the re-use of deep neural network features across data sets for the same tasks. We find that our method outperforms classical regression methods and training a network from scratch. In particular, we improve state-of-the-art results on age and sex prediction. Our transfer learning method may therefore provide a simple and efficient pipeline to achieve high performance on small MRI data sets.

## 1 Introduction

With the availability of big labeled data sets such as ImageNet [1], the application of deep neural networks [2] has shown huge success in a variety of fields such as computer vision, natural language processing [3] or speech [4]. In biomedical imaging, deep learning led to advances in areas such as automated diagnostics [5, 6, 7] and image segmentation [8], showing the huge potential of the application of artifical intelligence (AI) in healthcare [9]. In neuroimaging, the benefit of deep learning remains a subject of debate with some work showing that classical regression methods are not outperformed by deep learning techniques [10, 11, 12, 13, 14].

One challenge for applying deep learning in neuroimaging is that brain scans are usually expensive to acquire, which causes data sets to be small. On top of that, one faces the problem that there are significant differences in the image statistics across data sets, a phenomenon called *domain shift* (DS). In neuroimaging, this can be caused by technical differences in imaging protocols, image resolution, contrast, signal-to-noise ratio, or MRI hardware (e.g., MRI scanners at different hospitals) [15]. Moreover, DS occurs if different studies have different subject populations such as demographics or disease prevalence [16]. DS causes machine learning models which are trained on data from one dataset to perform significantly worse when applied to data from another dataset [17].

One approach to solve this is called *transfer learning* (TL). TL is a set of machine learning methods that enable to acquire knowledge from related tasks to improve performance on the task of interest - the *target task*. A doctor specialized in computed tomography (CT) could learn how to analyze MRI images much faster than someone with no background in radiology. TL tries to mimic such a transfer of knowledge on a machine learning level.

In this work, we try to solve the problem of DS in neuroimaging by the efficient re-use of features of deep neural networks across data sets. In computer vision, it is common practice to pre-train models on a large natural image dataset (e.g., ImageNet) and then finetune the models in the target small datasets to achieve faster convergence and better performance [18]. Yet, while several successful applications of deep learning in medicine such as skin cancer classification or detection of diabetic retinopathy follow this pipeline [19, 20], other work questions the benefit of such procedures in medical imaging [21].

In the last years, big brain imaging datasets such as UK Biobank (UKB) and the Human Connectome Project (HCP) have become available, opening new opportunities for harnessing the power of such data sets [22, 23]. Recently, these also enabled highly accurate machine learning in the neuroimaging domain. For example, Peng et al. [24] showed by proposing a light-weight deep neural network for 3d brain images that deep learning can outperform traditional machine learning techniques in brain age and sex prediction, on data from UKB. Here, we built on this work studying whether we can achieve high performance even for small data sets via transfer learning and whether a big neuroimaging data set such as UKB can help alleviate the problem of DS. To test our method, we use the three data sets from other studies: the OASIS-3 [25], ABIDE (I and II) [26] and IXI [27] data sets.

We focus our attention on age and sex prediction based on structual MRI data, as age and sex are labels which are available across most data sets. Predicting sex and age also serve as nontrivial surrogate tasks sharing challenges with tasks such as disease prediction. In addition, the prediction of age based on clinical MRI data is also clinically relevant as the difference between the predicted age and the actual age, i.e., the brain age delta, can serve as an effective biomarker [28], e.g., for predicting mortality [29].

We make the following contributions:

1. We systematically study the re-use of features of deep neural networks across scanners and show that features are re-usable even if DS occurs.
2. We propose an easy-to-use and effective transfer learning method to alleviate the problem of DS.
3. We show that via transfer learning, deep neural networks achieve significantly stronger performance than classical regression methods even in the small data regime. In addition, our results improve state-of-the-art on age and sex prediction.
4. We show that the superior performance can be attributed to the re-use of features learnt on the pre-training data set.

This paper is structured as follows. In section 2, the machine learning methods and data sets which we use are described in more detail. In section 3, we study the re-use of deep neural network features from where we derive our method and present experimental results. In section 4, we relate our results with previous work and discuss future directions.

## 2 Methods

### 2.1 Deep Learning

We begin by discussing the deep learning methods we use.

#### 2.1.1 Simple Fully Connected Neural Networks (SFCNs)

As a deep learning model, we use Simple Fully Connected Neural Networks (SFCNs) which were introduced by Peng et al. [24]. SFCNs built on the idea of fully convolutional neural networks [30]. Using almost exclusively convolutional layers as opposed to dense layers, the number of parameters of the deep neural network is drastically reduced allowing to build networks with many layers - even for the memory-intensive 3d MRI scans.

SFCNs consist of two main components: a feature extractor and a classifier (see fig. 1). The feature extractor receives a 3d MRI scan as input and outputs a high-dimensional vector. We interpret this as a *feature vector* of the scan following a common interpretation of deep neural networks [2]. Based on this feature vector, the classifier then outputs the predicted label.

**Figure 1:**
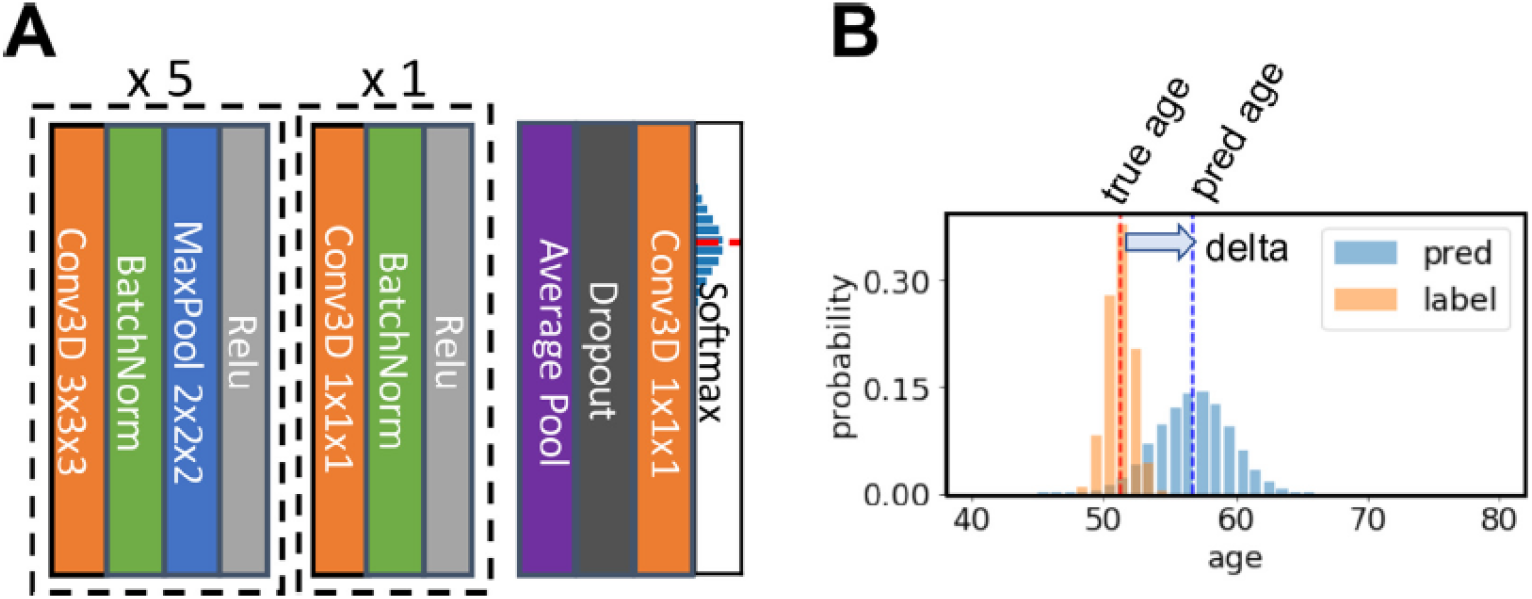
Illustration of SFCNs. **A:** the feature extractor consists of 6 blocks with convolutional layers, batchnorm, and Relu layers. The first 5 blocks have convolutions with kernel size 3 × 3 × 3 and an additional max-pooling layer, while the last block has kernel size 1 × 1 × 1. The classifier consists of an average pooling layer, a dropout layer, a fully connected layer, here similar to a 1 × 1 × 1 convolution, and a softmax layer. **B:** the output of SFCNs is a probability distribution over the possible labels. As depicted here for age prediction, the model is trained to output a discretization of the normal distribution 𝒩(*y*, 1) where *y* is the true age of the subject.

The feature extractor consists of 6 blocks: each of the first 5 blocks consist of a 3d convolutional layer of kernel size 3×3×3 with subsequent batch-normalization [31], max-pooling layer of size 2×2×2 and a final Relu non-linearity. The sixth block consists of a 3d convolutional layers of kernel size 1×1×1 with subsequent batch-normalization and a Relu non-linearity.

The classifier consists of an average pooling layer, a dropout layer [32], a 1 × 1 × 1 convolution, and a final softmax function giving a probability distribution *p =* (*p*_1_, …, *p*_*n*_) over *n* bins. If the model is trained on a classification problem, e.g., sex prediction, every bin would represent a class, e.g., female or male, and the predicted class would be the class of highest probability. If the model is trained on a regression problem such as age prediction, SFCNs frame this as a classification problems as well by assigning every bin *I =* 1, …, *n* a value range [*b*_*i*_, *b*_*i* + 1_]. The predicted age *ŷ* is then given by the expected value:

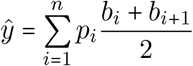

#### 2.1.2 Loss

To compute the loss between the output of the model, i.e., the probability distribution *p* = (*p*_1_, …, *p*_*n*_) across bins, and the true label *y*, we use two losses depending on the label type.

For age, we assign every age label *y* a bin distribution *q* = (*q*_1_, *q*_2_, …, *q*_*n*_) where *q*_*i*_ is the probability of bin [*b*_*i*_, *b*_*i*+1_] under the normal distribution 𝒩 (*y*, 1) with mean *y* and unit variance, i.e.

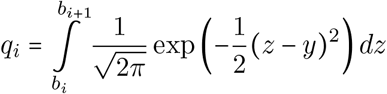

The loss is then the Kullback-Leibler divergence

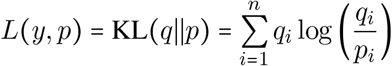

For sex, the model outputs a probability distribution *p* = (*p*_*f*_, *p*_*m*_) where *p*_*f*_ (resp. *p*_*m*_) is the probability for female (resp. male). We then use the cross-entropy as a loss between the true label *y* ∈ {*f, m*} and the output of the model:

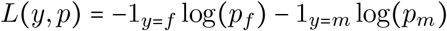

#### 2.1.3 Training

To train our model, we minimized the loss using Stochastic Gradient Descent (SGD). We trained our model using classical momentum of the learning rate [33]. We used a learning rate scheduler decaying the learning rate after *n* epochs by a factor of *λ* ∈ [0, 1].

An MRI scan is represented by an array of voxel intensities as input and we divide the array by its own mean intensity following previous work [24]. To increase the amount of data samples, we perform data augmentation in two ways (see fig. 2): first, we perform a random shift along the *x*-,*y*-, and *z*-axis by ±2 voxels. Second, we mirror an MRI scan with probability *p =* 0.5 along the sagittal subsection. This effectively increases the number of data samples by a factor of 250 and is therefore of particular importance since we target small data sets. We stress that this data augmentation is only performed during training and not during testing.

**Figure 2:**
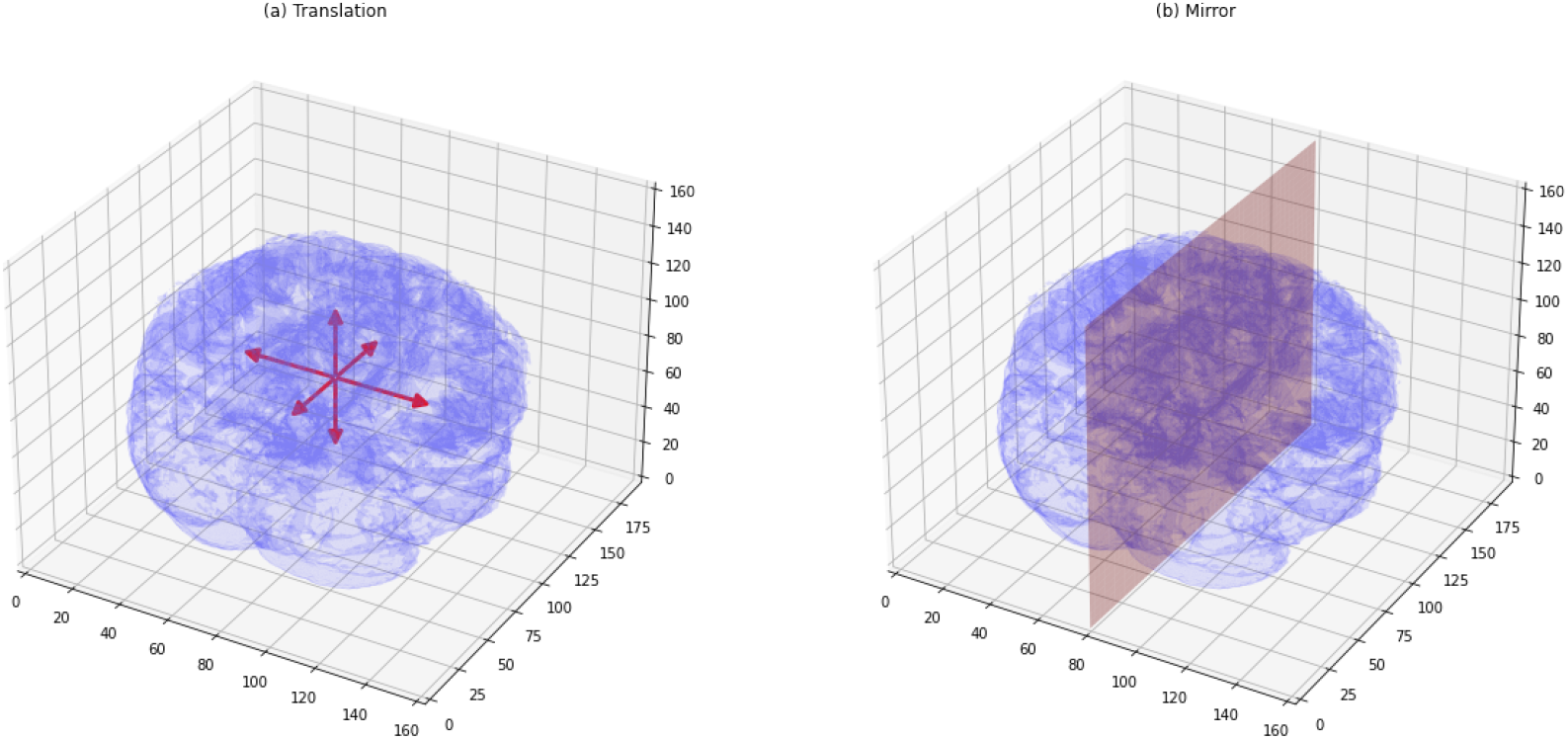
Illustration of data augmentation. (a) random translation along any of the three axis. (b) random mirroring along the sagittal plane

As common, we optimized the loss on the train set and optimized hyperparameters of the model on the validation set. The test set was untouched until all hyperparameters had been optimized.

#### 2.1.4 Implementation

To perform automatic differentation, we implemented SFCNs and the training procedures with *PyTorch* [34]. The models were trained on NVIDIA GeForce GTX 1080 Ti GPUs. Usually, computations were done with 2 GPUs in parallel. The github repository with the code can be found at: https://github.com/PeterHolderrieth/TransferLearning_Neuroimaging.

#### 2.1.5 Transfer Learning

In transfer learning [35], we are given two data sets which complement each other.

First, we are given a target data set 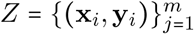 where (**x**_*i*_, **y**_*i*_) ∼ *P* are independent samples from a distribution *P*. We want our model to “learn” *P*. However, usually the size *m* of the data set is quite small. In our case, **x**_*i*_ corresponds to an MRI scan and **y**_*i*_ corresponds to age or sex.

Second, we are given a pre-training data set 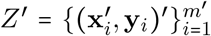 whose size *m*′ is usually big. However, there is a domain shift, i.e., the pairs 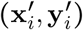 are samples 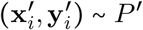 from a distribution *P*′ ≠ *P*, e.g., due to different properties of MRI scanners. Assuming that *P* and *P*′ are “similar”, the idea of transfer learning is to use the abundance of data from *P*′, i.e., the size *m*′ of *Z*′, to learn the task *P* of interest. In our case, *Z*′ is the UKB, and *Z* is one of ABIDE, IXI, or OASIS.

Our transfer learning pipeline is summarized in algorithm 1. One starts to randomly initialize the parameters of a model and trains it with the abundant data set *Z*′. In many cases, the output dimension, i.e., number of bins, may vary between *Z* and *Z*′. If this is the case, one first reshapes the classifier of the SFCN and re-trains the classifier keeping the feature extractor fixed. Finally, one trains the full model on the target data set *Z*.

To differentiate the training processes, we will use the term *pre-training* for training a randomly initialized model and the term *fine-tuning* for training a model whose parameters were loaded from a previous training process.

##### Algorithm 1: Transfer Learning with SFCNs

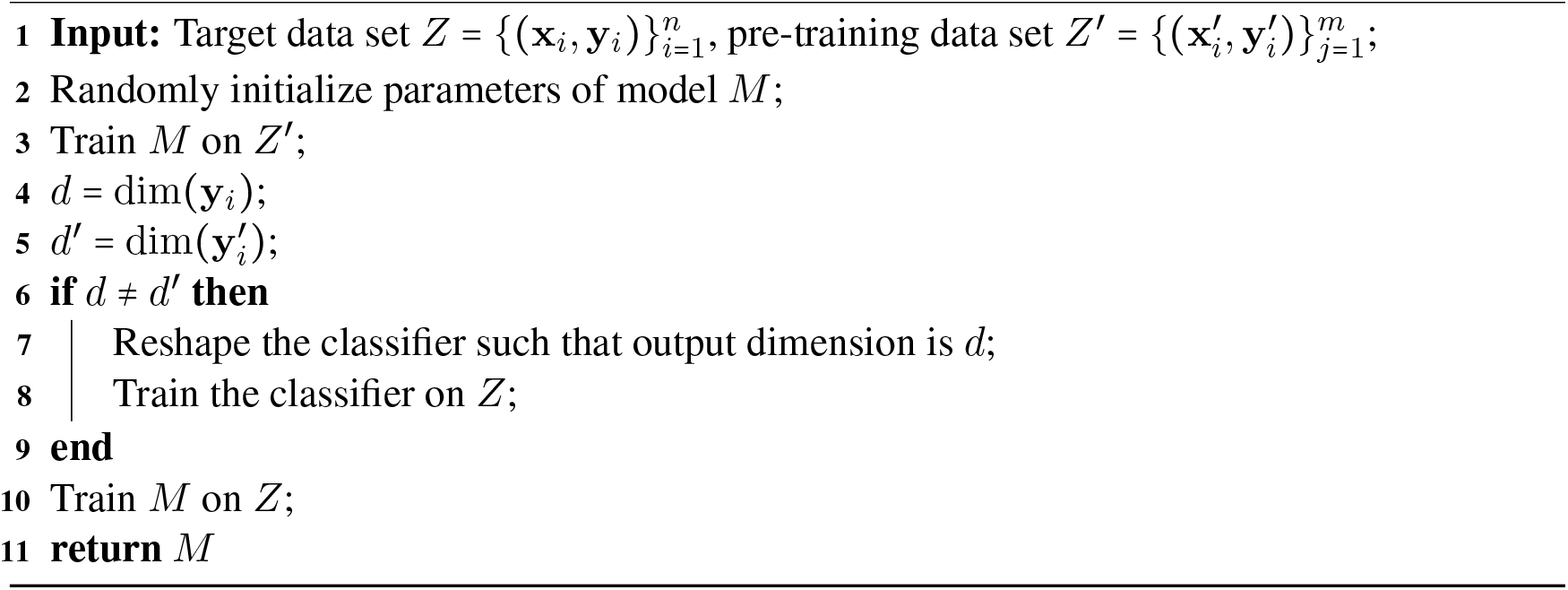

### 2.2 Data

#### 2.2.1 Data sets

##### UKB

As a pre-training data set, we use the UK Biobank (UKB). The UKB is a collection of brain imaging data from predominantly healthy participants [22]. In this work, we used the T1-weighted MRI scans from 40000 subjects. The image pre-processing pipeline is described in [36].

##### OASIS

The Open Access Series of Imaging Studies 3 (OASIS-3) [25] contains 1098 subjects and 2168 longitudinal MRI scanning sessions. Following LaMontagne et al. [25], the healthy control group consists of the subjects who have CDR score 0 from all clinical visits during the longitudinal studies. Only the 3T MRI sessions with valid clinical visit (within one year since or before the MRI scan) are included. This was because OASIS-3 contains both 1.5T and 3T MRI scans, while the CNN models were pretrained in UK Biobank scans using only 3T MRI. The selection process results in 1036 eligible subjects and 1806 eligible sessions.

##### IXI

The IXI dataset [27] contains MR images from normal, healthy subjects. We used the 3T scans resulting in 538 available scans.

##### ABIDE

The Autism Brain Imaging Data Exchange (ABIDE) initiative [26] collected MR images from both neurotypical and autistic subjects. We followed the selection process by [37].

In fig. 3, one can see various samples of MRI scans from the different data sets.

**Figure 3:**
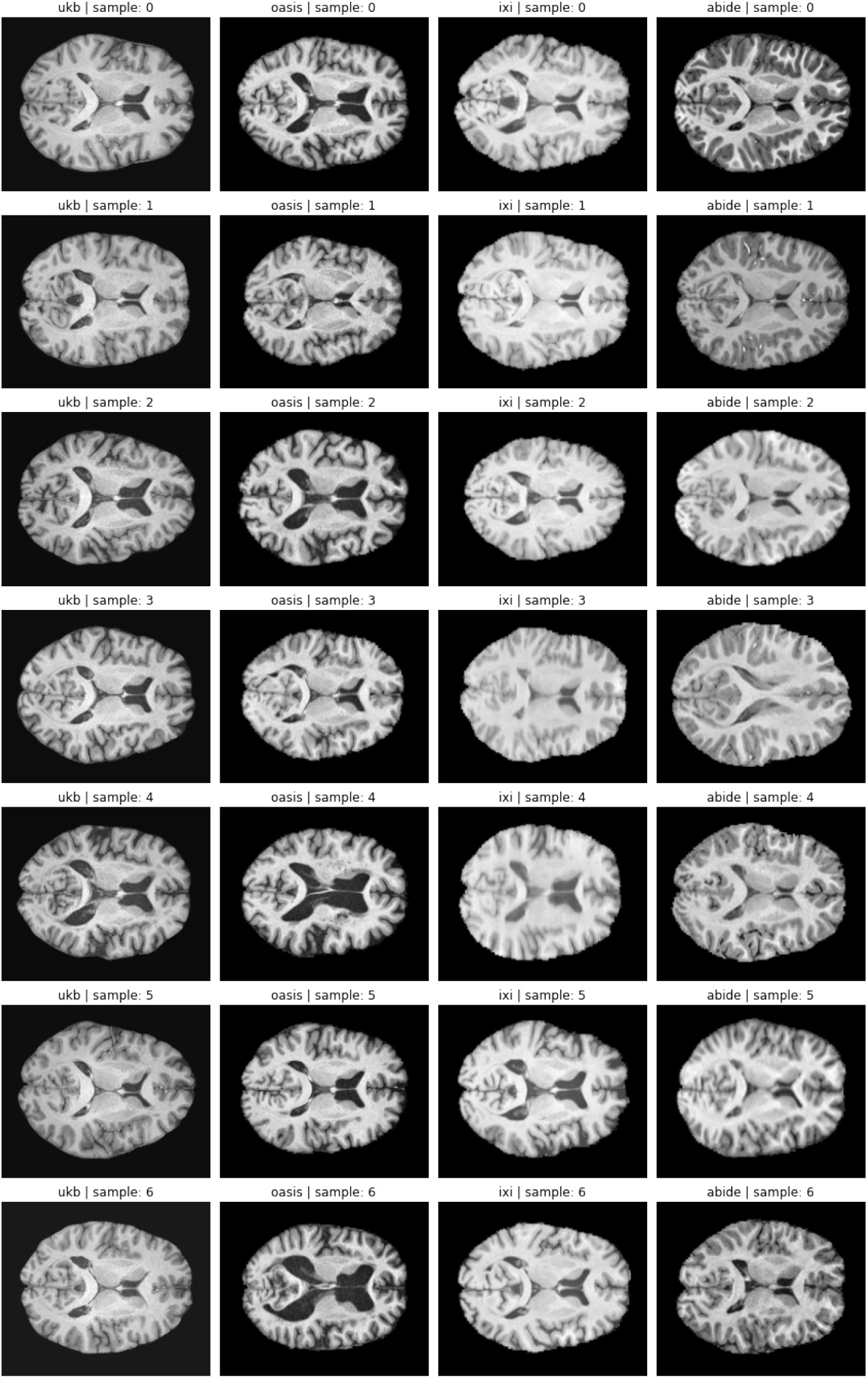
Samples of MRI scans (horizontal subsection). Every row gives a different sample. First column: UKB. Second: OASIS. Third: IXI. Fourth: ABIDE

#### 2.2.2 Data pre-processing

The preprocessing pipeline uses the *fsl anat* command from the FSL library [38] for automatic bias field removal, 12 degree of freedom linear registration, and non-linear registration to a standard space (2mm MNI 152 space), purely for the purpose of brain extraction. Finally, the 1mm brain is linearly transformed to MNI 152 with FLIRT [39, 40, 41] and used for the subsequent training/validation/testing for deep learning.

#### 2.2.3 Split of data sets

We split all data sets randomly in a train, validation, and test set (see table 1). The selection was done by randomly partitioning the subjects in a train, validation, and test group. For the training and validation set, we included all MRI sessions of a subject, while in the test set we only had one MRI session per subject.

**Table 1:** Summary statistics of data sets

|  | OASIS |  |  | IXI |  |  | ABIDE |  |  | UKB |
| --- | --- | --- | --- | --- | --- | --- | --- | --- | --- | --- |
|  | train | valid | test | train | valid | test | train | valid | test |  |
| *n* total | 355 | 91 | 686 | 215 | 108 | 215 | 537 | 269 | 537 | 39677 |
| *n* female | 228 | 50 | 272 | 120 | 63 | 114 | 147 | 70 | 127 | 20999 |
| *n* male | 127 | 41 | 414 | 95 | 45 | 101 | 389 | 199 | 409 | 18678 |
| age min | 42.7 | 48.4 | 45.8 | 20.1 | 20.2 | 20.0 | 5.2 | 5.43 | 5.32 | 44.6 |
| age max | 95.2 | 92.1 | 95.7 | 86.2 | 86.3 | 83.5 | 56.2 | 54.0 | 55.4 | 82.3 |
| age mean | 68.6 | 68.8 | 68.9 | 47.7 | 50.7 | 46.5 | 13.6 | 13.6 | 13.2 | 64.1 |
| age std | 8.1 | 9.7 | 8.8 | 16.7 | 16.7 | 15.2 | 6.52 | 6.2 | 6.0 | 7.5 |

In OASIS, we performed a completely random split. In IXI and ABIDE, the sampling process considered scanner site, proportion of females, and the age distribution, ensuring a similar distribution per respective class in every train, validation, and test set. As one can see in table 1, the numbers of samples per data set is significantly smaller than the UKB and the proportion of females varies significantly across OASIS, IXI, and ABIDE. Moreover, the age distribution varies as well, e.g., ABIDE covers relatively young subjects of 5-55 years, while OASIS covers an older population of 42-95 years. In sum, these data sets represents a variety of realistic domain shifts in terms of subject population and serve as good examination tasks for transfer learning.

As one can see in table 1, the number of female and male subjects vary across data sets. If we train or test models on sex prediction, we therefore balanced the data sets by oversampling the underrepresented sex, i.e., we randomly sampled subjects from the underrepresented sex and inserted them into the data set.

On top of subject population, we would also expect a significant domain shift in terms of technical difference across scanning procedures. As one can see in table 2, the number of different scanner models used and the number of different scanner sites vary significantly. These differences might also explain the variations seen in fig. 3: OASIS and UKB seem relatively similar in contrast and brightness, while IXI seems brighter and ABIDE shows big variation in terms of brightness.

**Table 2:** Technical acquisition details about data sets

| Data | Scanner brands | $n$ scanner models | $n$ sites |
| --- | --- | --- | --- |
| UKB | Siemens | 1 | 3 |
| OASIS | Siemens | 1 | 2 |
| IXI | Philips, GE | 3 | 3 |
| ABIDE | Philips, GE, Siemens | 6 | 19 |

### 2.3 Elastic net regression

To perform regression on the MRI scans, we perform principal component analysis (PCA) on the train data and choose the *k* principal components which are most correlated with the labels (either age or sex). We then perform elastic net regression [42] on these principal components. To compute predictions based on the validation and test data, we simply project the data to the *k* previously selected features and perform predictions with the fitted linear regression parameters.

## 3 Results

### 3.1 Transfering deep neural networks across MRI scanners

We first study the effects of using a deep neural network for MRI scanner sites or data sets which it was not trained on. These results give a justification for our transfer learning method (see algorithm 1) and empirically show the effects of domain shift.

#### 3.1.1 Direct transfer

We begin by studying the change in performance after domain shift. We look at sex prediction and use 5 models which were pre-trained on the UKB with different random seeds. On UKB data, we achieve very accurate results of 99.5%. However, as one can see in table 3, there is a significant decline in performance when testing such a model on different data sets. The high performance on OASIS might be explained by the similarities of age distribution and scanners to the UKB (see section 2.2.3 and fig. 3). While performance varies significantly with accuracy of 90.2% on OASIS and 59.1% on IXI, in all data sets we observe a significantly higher performance than random selection, i.e., higher than 50%. This indicates that pre-training on UKB equips a model with an inference rule which is better than random initialization and at least partially transferable across data sets. It seems therefore sensible to re-use the information from a model even if it was trained on a different data set.

**Table 3:** Testing SFCNs pre-trained on UKB sex classification on different data sets. Mean and standard deviation over 5 runs. Data sets were balanced for sex.

| Dataset | Accuracy |
| --- | --- |
| UKB | 99.5% ± 0.1% |
| OASIS | 90.2% ± 2.5% |
| ABIDE | 59.9% ± 4.9% |
| IXI | 59.1% ± 7.0% |

#### 3.1.2 Feature re-use and task specificity of features

We next study why such a decline in performance occurs. As explained in section 2.1.1, SFCNs consist of a feature extractor and a classifier. We can visualize how SFCNs learn to extract information from MRI scans by visualizing the high-dimensional features produced by the feature extractor (see figs. 4 and 5).

**Figure 4:**
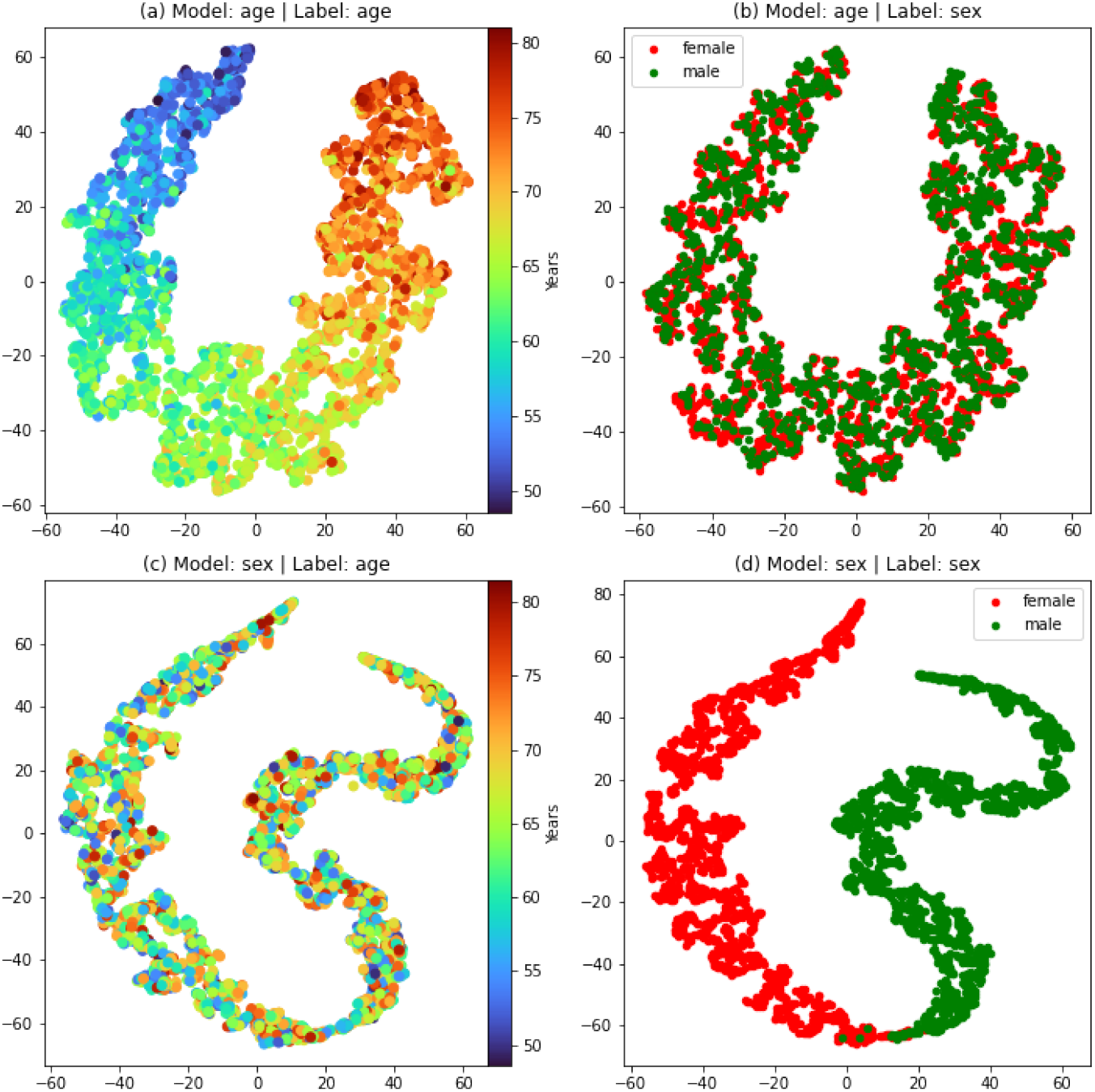
Plotting SFCN features of UKB via t-SNE. (a),(b): model trained on age. (c),(d): model trained on sex. While model learn a meaningful representation of the features, one can observe that the features are highly task-specific, i.e., a model trained on age does not spatially differentiate sex and a model trained on sex does not separate age.

**Figure 5:**
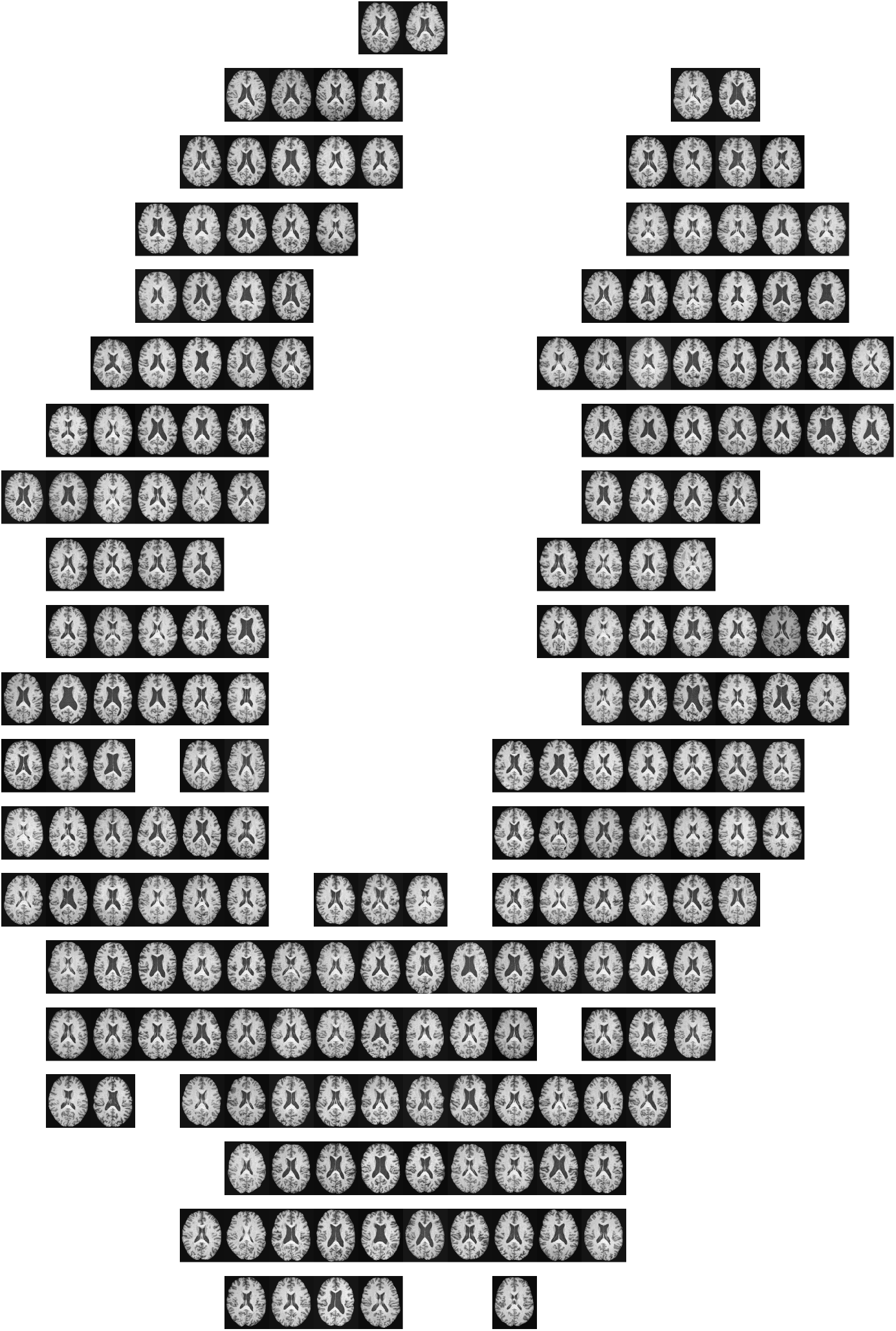
Illustration of t-SNE plot (fig. 4a) with “brain manifold”

We use two models which were trained on the UKB. The first one was trained on age prediction and the second one was trained on sex prediction. Passing MRI scans from the UKB test set through the feature extractor of the pre-trained models, we perform dimensionality reduction on the acquired data via t-distributed stochastic neighborhood selection (t-SNE) [43] (see fig. 5 as an illustration). As one can see in fig. 4, the feature extractor spatially separates MRI scans in a “manifold” giving a continuous spectrum of age and a distinction between female and male participants. It is particularly striking that features are highly specific to the task which the model was trained on. For example, the features from a model trained on age prediction do not spatially separate scans of female and male subjects (see fig. 4 (b)). It therefore seems plausible that for transfer learning, we should use models which were pre-trained on the same task. Next, we visualize how domain shifts affect the feature extraction (see fig. 6). Here, we only visualize the labels which the model was pre-trained on the UKB to predict. One can observe that in OASIS and IXI, the spatial separation of age and sex is highly preserved (see fig. 6 (a,b,d,e)). In contrast, features extracted from the ABIDE data show a high degree of confusion.

**Figure 6:**
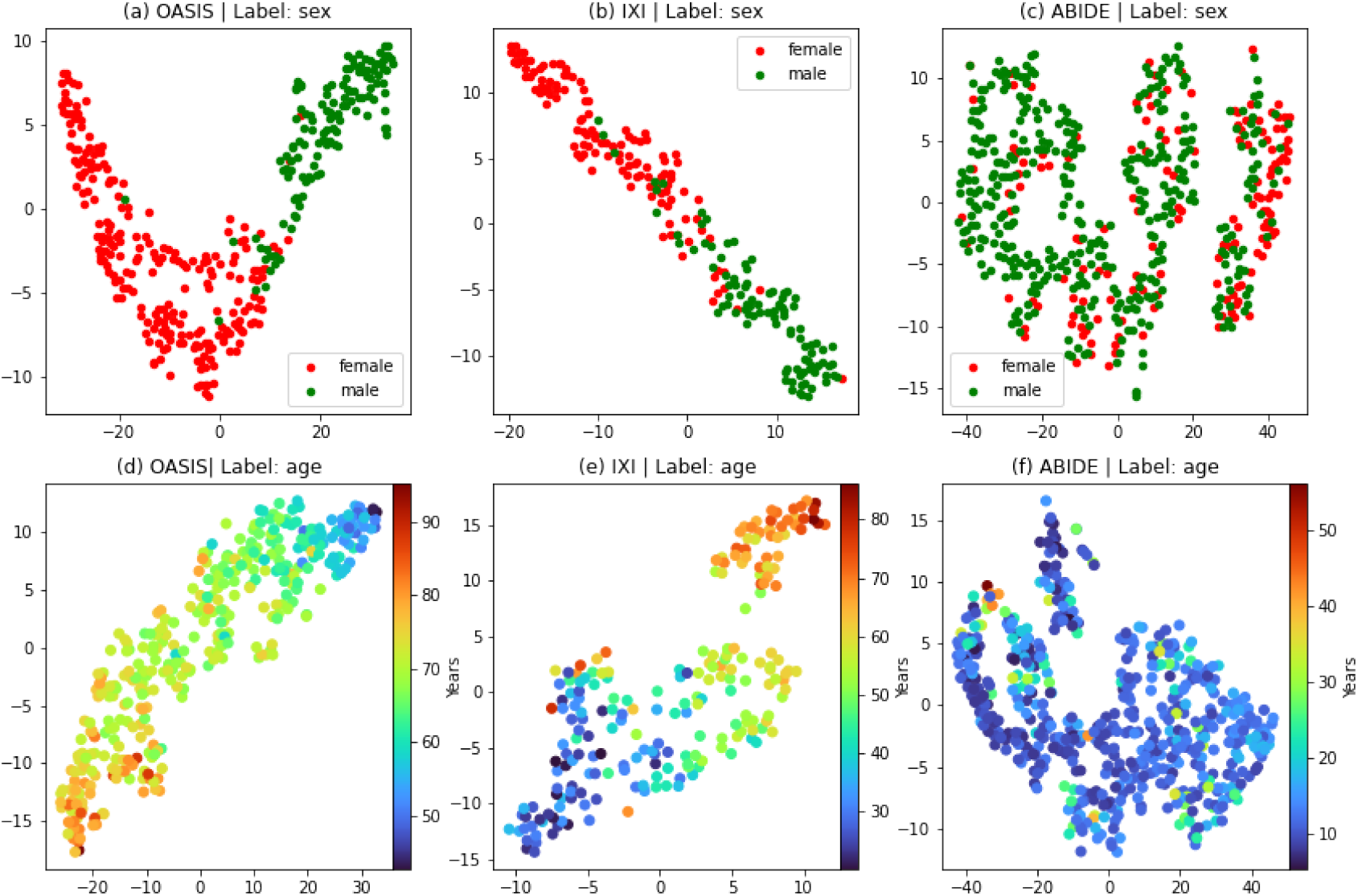
t-SNE plot of SFCN features after domain shift from UKB to various data sets. (a),(b),(c) model pre-trained on UKB sex prediction. (d),(e),(f) model pre-trained on UKB age prediction.

It is known that the MRI patterns of aging are completely different in the sexes and across ages [44]. Similarly, sex differentiation varies across ages. This might explain the high degree of confusion in the features for ABIDE: ABIDE has an age range 5 − 56 of compared to an age range of 44 − 82 of the UKB (see tables 1 and 2). A model pre-trained on the UKB was trained to learn age and sex differentation patterns for subjects older than subjects in ABIDE. For ABIDE and UKB, the orthogonal age distributions of their respective subject populations make pre-trained features less useful.

To quantify the effect of domain shifts on the features of a deep neural network, we fine-tune the parameters of the classifier of a pre-trained model on the target set while keeping the feature extractor constant (steps 1-9 in algorithm 1). Crucially, we use feature extractors from models pre-trained on different tasks. This allows us to evaluate how important the task is where the features were pre-trained on. In table 4, one can see that the models which were pre-trained on the same task outperformed models which were pre-trained on a different task. This confirms the observation from fig. 4 that features are task-specific. Moreover, the highly accurate results confirm the observation from fig. 6 that features can be highly re-usable across data sets if they are from the same task.

**Table 4:** Results on age or sex prediction on OASIS by fine-tuning the classifier of models which were pre-trained on the UKB on age or sex prediction (mean and std over 5 runs). For sex prediction, the data sets were balanced for sex. “MAE” is mean absolute error. “Acc.” is accuracy.

| Pre-trained (UKB) | Task (OASIS) | Results |
| --- | --- | --- |
| Age | Age | $3.52 \pm 0.02$ (MAE) |
| Sex | Age | $6.91 \pm 0.01$ (MAE) |
| Sex | Sex | $94.6\% \pm 0.2\%$ (acc.) |
| Age | Sex | $52.0\% \pm 0.5\%$ (acc.) |

#### 3.1.3 Feature fine-tuning

So far, we have shown that steps 1-9 in algorithm 1 significantly improves performance. We finally study to which extent it is beneficial for model performance to perform step 10, i.e., to fine-tune the feature extractor. In principle, there should be no decrease in performance since picking a learning rate *λ =* 0 would simply mean leaving out this step. However, it is possible to overfit by picking hyperparameters optimizing results on the validation set.

In table 5, we compare the results between the full algorithm or fine-tuning only the classifier. As one would expect, we see a consistent improvement in performance on the validation set for all data sets apart from ABIDE for sex prediction. However, the test results are less clear. While we can see some overfitting in OASIS sex prediction and IXI age prediction, fine-tuning the full model usually improves results. In particular, we can see a particular big improvement for ABIDE. This fits to observation from fig. 6 showing a high confusion of features for ABIDE making feature fine-tuning more necessary as opposed to OASIS where features are more ordered from the start and overfitting is more prominent.

**Table 5:** Comparing results between fine-tuning the classifier and algorithm 1. For sex prediction, the data sets were balanced for sex. “MAE” is mean absolute error. “Acc.” is accuracy. “Valid” refers to results on the validation set, “test” to results on the test set.

| Data | Task | Method | Results (valid) | Results (test) |
| --- | --- | --- | --- | --- |
| OASIS | Age | ft. class | $2.96 \pm 0.05$ (MAE) | $3.52 \pm 0.02$ (MAE) |
| | | algorithm 1 | $2.82 \pm 0.05$ (MAE) | $3.25 \pm 0.02$ (MAE) |
| OASIS | Sex | ft. class | $90.8\% \pm 0.7\%$ (acc.) | $94.6\% \pm 0.2\%$ (acc.) |
| | | algorithm 1 | $97.0\% \pm 1.6\%$ (acc.) | $92.6\% \pm 1.8\%$ (acc.) |
| IXI | Age | ft. class | $6.12 \pm 0.07$ (MAE) | $5.35 \pm 0.10$ (MAE) |
| | | algorithm 1 | $5.82 \pm 0.59$ (MAE) | $5.65 \pm 0.55$ (MAE) |
| IXI | Sex | ft. class | $80.0\% \pm 0.4\%$ (acc.) | $87.6\% \pm 0.3\%$ (acc.) |
| | | algorithm 1 | $85.6\% \pm 2.6\%$ (acc.) | $90.3\% \pm 1.3\%$ (acc.) |
| ABIDE | Age | ft. class | $4.39 \pm 0.21$ (MAE) | $4.43 \pm 0.22$ (MAE) |
| | | algorithm 1 | $1.72 \pm 0.06$ (MAE) | $1.70 \pm 0.07$ (MAE) |
| ABIDE | Sex | ft. class | $74.4\% \pm 0.6\%$ (acc.) | $70.2\% \pm 2.3\%$ (acc.) |
| | | algorithm 1 | $69.7\% \pm 5.4\%$ (acc.) | $75.7\% \pm 4.5\%$ (acc.) |

In fig. 7, we visualize the change of feature extraction applying algorithm 1 for ABIDE age prediction and OASIS age prediction. As one would expect from the results of table 5, one can see a clear evolution of an age spectrum for ABIDE, while OASIS features only change little since the already separated the ages from the start.

**Figure 7:**
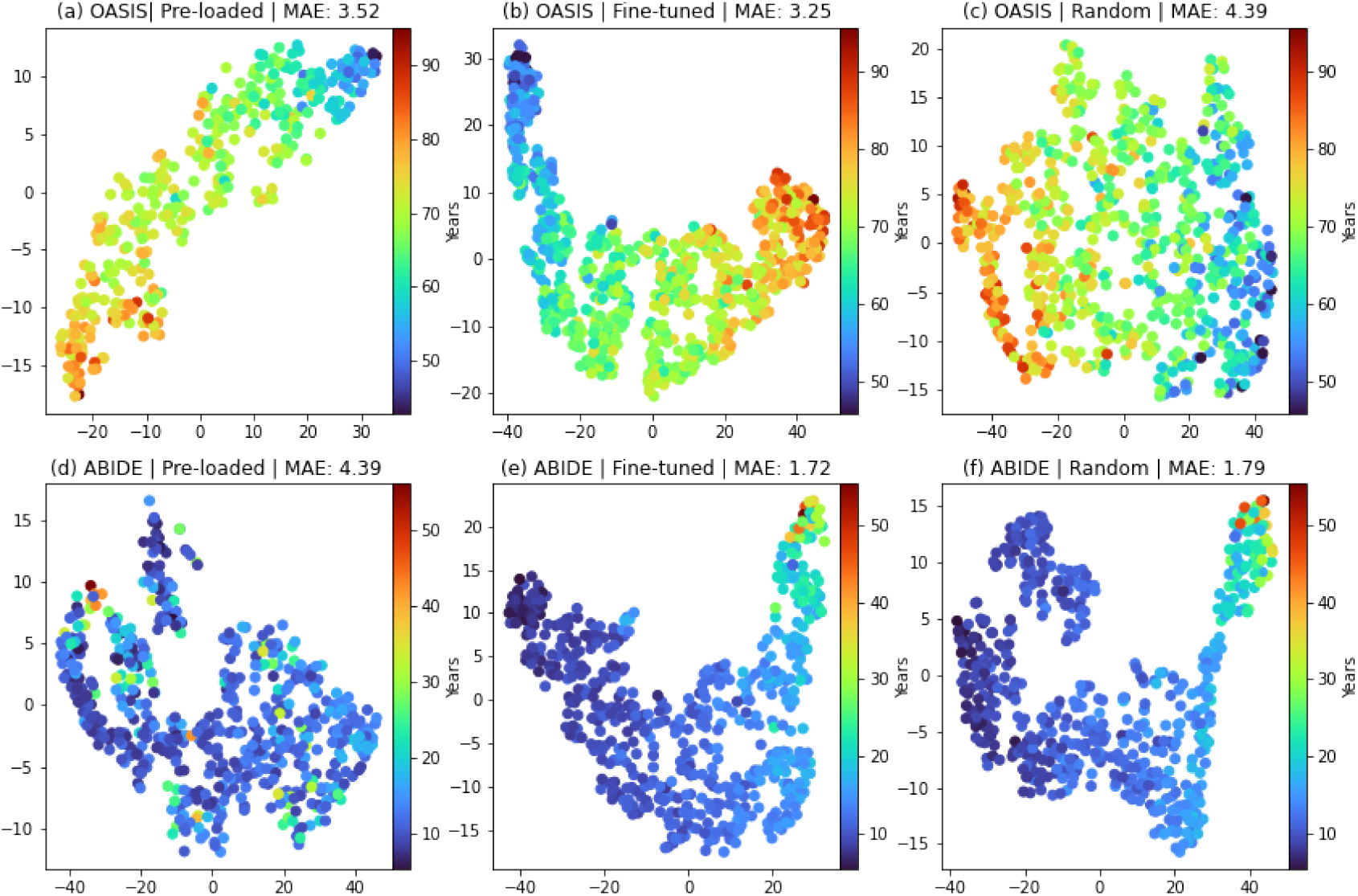
Plotting SFCN features after fine-tuning the feature extractor and training a model from scratch. (a,d) Features of OASIS/ABIDE (test set) based on UKB age prediction model. (b,e) After fine-tuning the feature extractor on OASIS/ABIDE (train set). (c,f) Features of ABIDE (test set) based on UKB age prediction model. For OASIS, the features are re-usable (a), improve by fine-tuning (b) and are not reproducible by training from scratch (c). For ABIDE, pre-trained features are not useful (d) and only fine-tuning the feature extractor brings performance gains (e), while training from scratch can create similar features (f).

Together, this subsection shows that algorithm 1 successfully exploits the re-use of deep neural networks across data sets and represents a sensible transfer learning pipeline for neuroimaging.

### 3.2 Transfer learning vs. classical regression methods

We also compare our transfer learning pipeline (algorithm 1) with a classical regression method as described in section 2.3. Our results in table 6 show that the transfer learning pipeline outperforms regression methods across data sets and across tasks by a large margin. For age prediction, the mean absolute error (MAE) is approximately halved by using transfer learning compared to elastic net regression. For sex prediction, the difference of the accuracy to 50% (i.e., random selection) is doubled for transfer learning compared to regression.

**Table 6:** Comparing elastic net regression (see section 2.3) with transfer learning (see yalgorithm 1). For age prediction, naive prediction is prediction of the mean of the training set. For sex prediction, it is random selection. For sex prediction, the data sets were balanced for sex.

| Dataset | Method | Age | Sex |
| --- | --- | --- | --- |
| OASIS | Naive | $6.94 \pm 0.00$ | $50.0\% \pm 0.0\%$ |
| | Elastic | $6.24 \pm 0.52$ | $67.0\% \pm 3.9\%$ |
| | Algorithm 1 | $3.25 \pm 0.02$ | $92.6\% \pm 1.8\%$ |
| IXI | Naive | $13.16 \pm 0.00$ | $50.0\% \pm 0.0\%$ |
| | Elastic | $9.90 \pm 0.26$ | $73.1\% \pm 4.7\%$ |
| | Algorithm 1 | $5.65 \pm 0.55$ | $90.3\% \pm 1.3\%$ |
| ABIDE | Naive | $4.12 \pm 0.00$ | $50.0\% \pm 0.0\%$ |
| | Elastic | $3.59 \pm 0.60$ | $66.9\% \pm 1.8\%$ |
| | Algorithm 1 | $1.70 \pm 0.07$ | $75.7\% \pm 4.5\%$ |

### 3.3 Comparing results to other works

In general, it is difficult to compare prediction results to previous work: train, validation, and test sets have different sizes and different choice of modalities and pre-processing methods might affect performance. However, we name a few to put our results into context.

For OASIS, Dinsdale et al. [45] perform age prediction on OASIS with a training size of 813 (compared to 355 here). We improve the result from 3.79 MAE to 3.25 MAE, although we use less training data.

Jónsson et al. [46] use IXI for age prediction. They link the the difference between predicted and actual gene with two genomic sequence variants correlating with reduced sulcal width and reduced white matter surface area. Their result on IXI is 4.149 MAE compared to 5. in our work. However, the size of their training data set is 440 compared to 215 in this work. Most importantly, they used IXI for *validation*, i.e., they optimized hyperparameters on IXI and they did not report a test result on IXI. Therefore, a direct comparison to our work is not possible.

For ABIDE, Bellantuono et al. [37] achieve an MAE of 2.19 on ABIDE age prediction (compared to 1.72 here) using a larger test set and excluding 22 subjects older than 40. Our method therefore clearly improves state-of-the-art results on ABIDE age prediction.

### 3.4 Explaining the performance gain of transfer learning

To study the high performance of transfer learning, we next compare our method with training a model from scratch, i.e., randomly initialize the weights and biases of the network instead of loading them from a pre-trained model. As one can see in table 7, transfer learning outperforms training from scratch in all cases apart from ABIDE sex prediction. As one might expect from fig. 6, the pre-trained features for ABIDE sex prediction are not useful and even lead to worse results than training from scratch. This phenomenon is called *negative transfer* which refers to reduced accuracy due to transfer learning mainly caused by the source and target domain not related “enough” [47]. In the context of MR brain imaging, this has already been observed to lead to worse results [48].

**Table 7:**
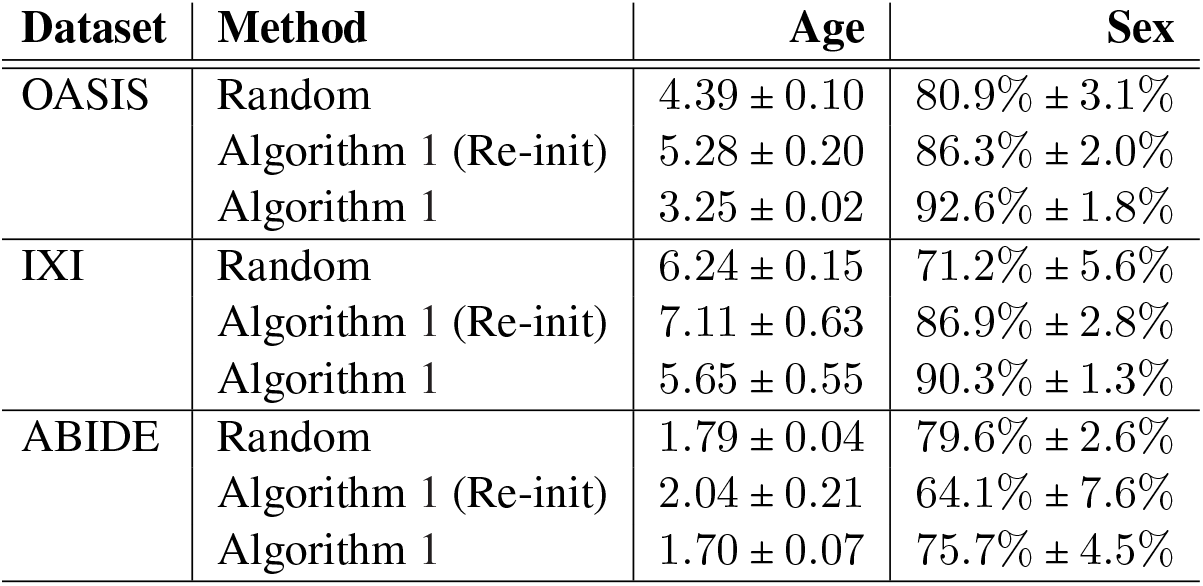
Comparing transfer learning with random initialization of parameters of the network. “Random” denotes training from random initialization via classical SGD. Algorithm 1 (Re-init) follows algorithm 1 but before step 4 randomly re-intializes every layer of the model *M* with mean and variances given by the pre-trained parameters.

In previous work, it was claimed that the performance gain of transfer learning can mainly be attributed to the correct scaling of parameters of the networks as opposed to actual re-use of features [21]. To test this, we compare the performance of transfer learning with the following method of random initialization of weights: every layer is randomly initialized with mean and variance given by the mean and variance of the weights of a pre-loaded model. Crucially, we use algorithm 1 to train this model (not training from scratch). The results in table 7 show that this scaled random initialization does not show a performance comparable to algorithm 1. This shows that it is the actual feature transfer or feature re-use which explains the boost in performance due to transfer learning.

Overall, the results in table 7 show that it is not merely using a deep neural network but transfer learning which brings meaningful performance gains.

### 3.5 Transfer learning and data scarcity

The scarcity of data is one of the main challenges in applying deep learning to neuroimaging. For a successful application of our transfer learning pipeline, it is therefore crucial how many data samples are needed to improve accuracy. We study this here. Beyond the transfer learning pipeline, we also test fine-tuning only the classifier (i.e., final layer) of the model. We do experiments varying the size of the training set keeping the size of the test set constant.

In fig. 8, we can see that for OASIS age prediction, the full transfer learning algorithm gives very accurate results even for as few training samples as 50. It also consistently outperforms fine-tuning only the final layer. In contrast, for ABIDE sex prediction, we observe that algorithm 1 is only better if the number of training samples is higher than 400. With too few samples, we observe that the full transfer learning pipeline overfits to the small training set. Therefore, only fine-tuning the final layer might be a more sensible choice in the very small data regime.

**Figure 8:**
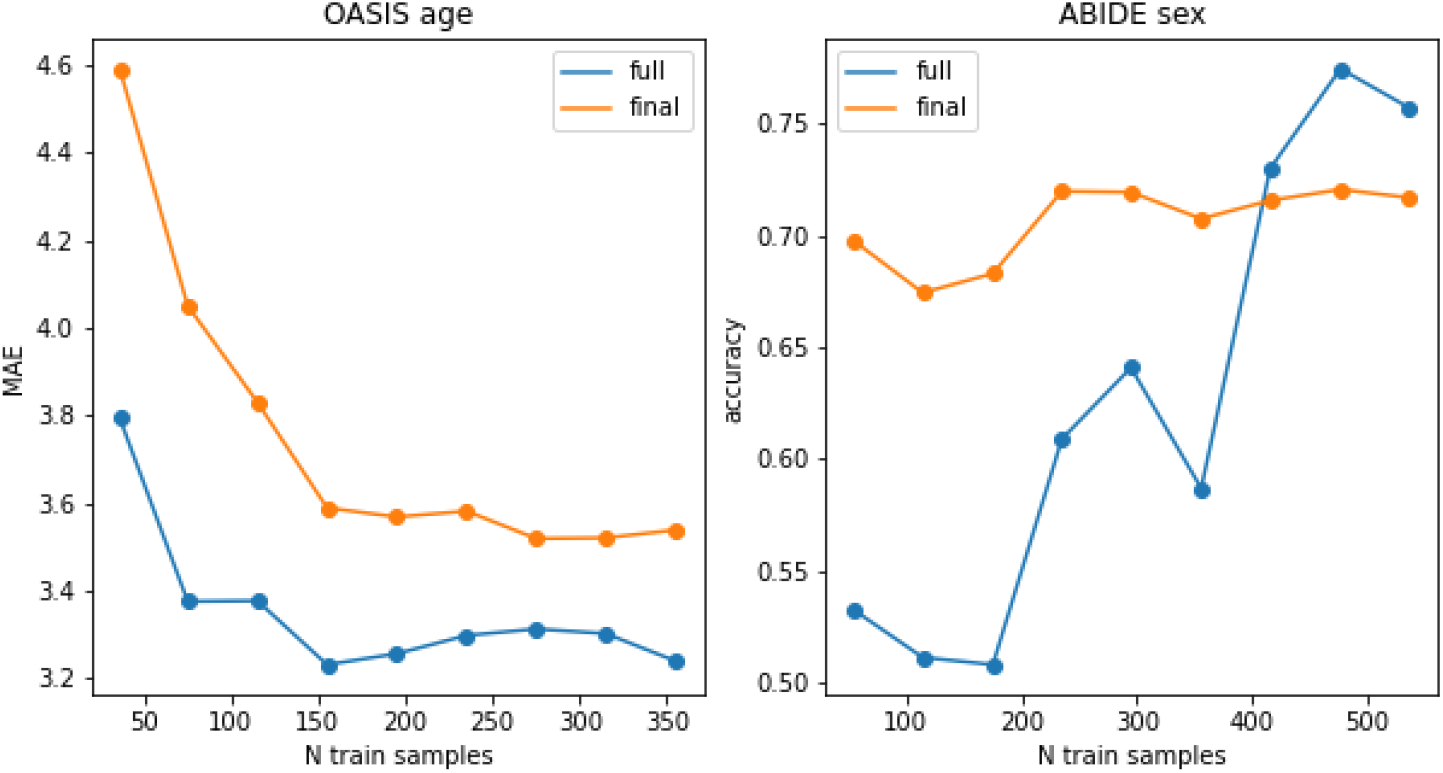
Studying performance depending on the size of the training set. ‘full’ refers to the full transfer learning method (algorithm 1), and ‘final’ for fine-tuning only the final layer. Left: OASIS age prediction. Right: ABIDE sex prediction.

### 3.6 Convergence time

Beyond performance accuracy, there are other practical advantages which transfer learning might bring. Intuitively, a pre-trained model has a “headstart” and needs less training iterations. We study this for sex prediction on OASIS, IXI, and ABIDE.^1^ In fig. 9, we can indeed observe a stronger convergence. Even in cases where the performance gain of transfer learning is minor such as for ABIDE, one can observe a clear speed-up of training due to transfer learning. Hence, transfer learning is beneficial not only for performance but also for training efficiency.

**Figure 9:**
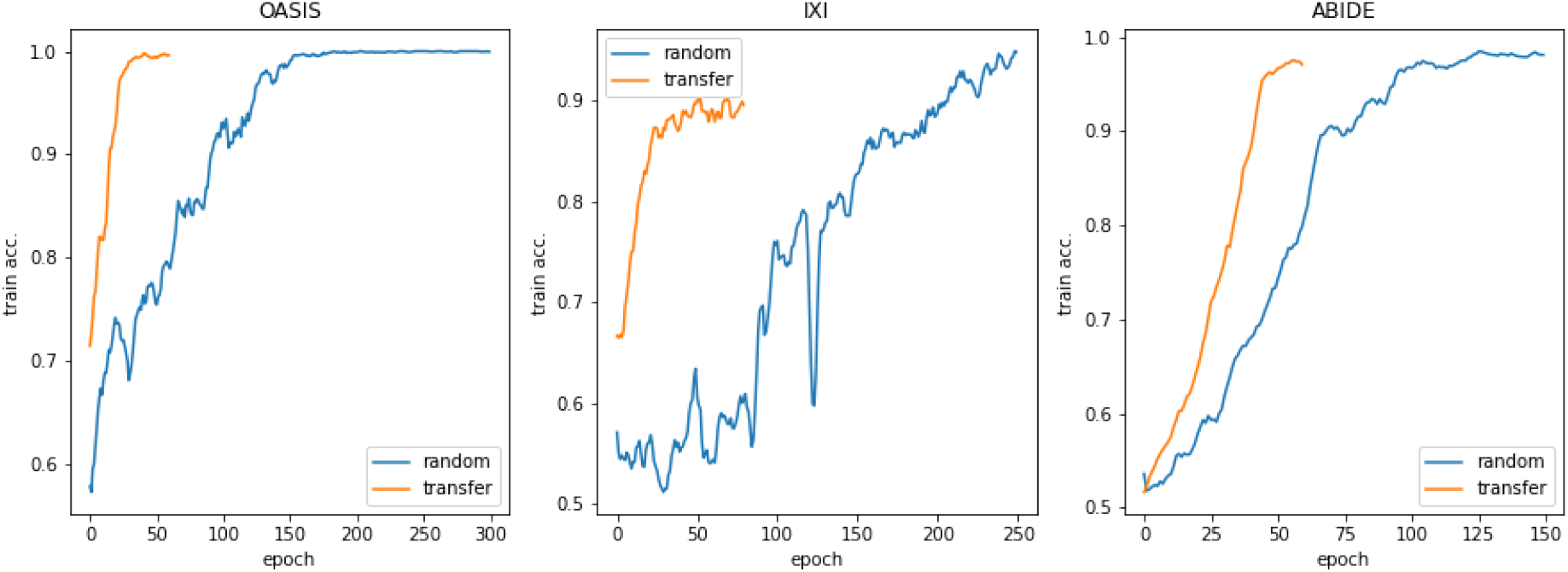
Comparing training convergence between randomly initialized models and transfer learning. One can see that the training accuracy improves much quicker and flattens much faster for a pre-trained model.

## 4 Discussion

### 4.1 Pre-training on large data sets

In this work, we showed that “simple” transfer learning leads to performance improvements for neuroimaging as it does for natural image data sets. This is particularly interesting when comparing our work to Raghu et al. [21], who argue that fine-tuning pre-trained models does not bring any performance gains in biomedical imaging. However, our work is not inconsistent with their results since we use a neuroimaging data set for pre-training as opposed to a natural image data set. Rather, we have shown (see section 3.1.2) that features which a neural network learns from data are very specific to the neuroimaging domain, even specific to the tasks such as age and sex. Similarly, visualizations of convolutional kernels trained on ImageNet have been shown to lead to features that are suited for detecting natural images [49]. As Raghu et al. [21] also remarked, it therefore seems plausible that pre-training on a natural image data set such as ImageNet does not bring any performance gains in the biomedical domain beyond favourable initial scaling of the weights, while data sets such as UKB do.

In computer vision, the discovery that features learned on a pre-training task can be useful to a target task [50, 49] have led to the following practice: initial pre-training models on ImageNet with subsequent fine-tuning on the target task. From a computer vision perspective, we showed in this work that the UKB might serve as the “ImageNet” for neuroimaging, i.e., UKB can provide a training task for a deep neural network which is “similar enough” to the task of interest. While we used the currently available ∼ 40000 samples, the UKB continues to grow in size up to 100000 in the next few years [22]. Its power to enable effective and simple transfer learning is therefore likely to grow as well.

However, there are certain challenges. In neuroimaging, technical variations have been observed to be a much more significant problem than for natural images, and UKB does not cover the whole spectrum of MRI scanners, image protocols, etc. (see table 2). More importantly, since the UKB is not disease-specific, it is difficult to pre-train a model on disease prediction. Given the results in table 4, it therefore remains an open question how to transfer across tasks and how to use the UKB for direct disease prediction as opposed to biomarkers such as the brain age delta.

### 4.2 Traditional machine learning vs. deep learning

While the advantages of deep learning above regression methods have been a subject of debate, recent work showed that deep learning outperforms traditional regression methods for age and sex prediction on UKB [24].

Considering that our method mainly relies on re-training the classifier, it can be considered as a regression method based on features previously learnt from a big neuroimaging data set. Therefore, the claim that regression methods outperform deep learning on small data sets might still be true - we simply use more informative features. This is particular striking given results in fig. 8 which show that fine-tuning the final layer (pure regression on the features) can outperform the full transfer learning method if there are only few data samples available.

However, comparing table 6 and table 7 we observe that even random initialization outperforms regression methods. Therefore, our work shows that the superior performance of deep learning also applies to the small data regime - without but even more significantly with transfer learning.

### 4.3 Transfer learning vs. training from scratch

For natural images and computer vision, He et al. [51] challenge the assumption that pre-training on ImageNet brings performance gains beyond training from scratch. They report similar results of transferred and randomly initialized models. However, they find that transfer learning leads to faster convergence. In this work, we also show that transfer learning speeds up convergence. But in contrast, we show that for neuroimaging transfer learning actually brings performance gains in terms of accuracy.

### 4.4 Comparison with other transfer learning approaches

#### 4.4.1 Feature-based approaches

While our transfer learning method also fine-tunes the full model, we showed that the main performance gains come from fine-tuning the classifier, i.e., the parameters of the final layer of our network, for every target data set (see table 5). However, other works are more “feature-based” in the sense that transfer learning happens on a feature level - for SFCNs this would be the feature extractor (see fig. 1).

Symmetric feature-based approaches harmonize features from the source and target domain. For example, Dinsdale et al. [45] proposed a method based on adversarial training, unlearning information about the scanner sites, enabling multi-scanner integration and generalization with a *single* classifier across data sets. Other works performed such an harmonization on the feature level by minimizing their distances via different metrics such as mutual information [52] or maximizing their correlation [53, 54]. The results of this work show that a harmonization on the feature level is not necessarily needed if the differences across data sets are small (e.g., OASIS and UKB, see fig. 6 and table 2). However, it is interesting that the “most inhomogeneous” data set, i.e., ABIDE with its big variety of scanner models used (see table 2), led to the worst results for us. Therefore, the need for harmonizing different data sets on a feature level seems to remain and it would be interesting to study a combination of a “feature-based” and a “classifier-based” approach. However, such approaches would require to re-train a model for every new small “target” dataset - a challenge in applications.

#### 4.4.2 Style transfer approaches

A further approach to alleviate the problem of domain shift is using style transfer methods. Such methods allow to transfer an MRI scan of “style A”, e.g., the image properties of MRI scanner A, to “style B”, e.g., the image properties of MRI scanner B [55]. Ali et al. [56] propose a transfer learning method based on Generative Adversarial Network (CycleGAN) [57] to transfer images across different sites and provide promising results. Similarly, Bashyam et al. [58] use CycleGANs to map MRI data from different data sets to a canonical reference domain where the image appearance does not depend on the scanner site.

A style-transfer approach could also extend our transfer learning method: instead of using UKB data for pre-training (step 3 in algorithm 1), one could create an OASIS-style UKB (resp. ABIDE-style, IXI-style UKB) via CycleGANs and use it for pre-training. Subsequently, one could continue with algorithm 1 as above. Future work could explore whether such an approach improves results. In this work, we focused on using simple methods since these promise to be of greatest use in the clinic.

#### 4.4.3 Instance-based approaches

Another possibility to solve the problem of domain shift is to take an *instance*-based approach where similarities of the MR images themselves are modelled to transfer knowledge. For example, previous work weighted test images based on the similarity to the training images and classifying via weighted Support Vector Machines (SVMs) [59]. Cheng et al. [60] performed a similar analysis with combined feature selection and sample selection. Unfortunately, in our case such an approach would mean re-training the model for every test which is impractical with deep learning methods and the current computational power.

#### 4.4.4 Methods different from transfer learning

There are various related strategies to transfer learning: first, meta-learning algorithms *train* models to learn from less data. For example, Liu et al. [61] showed that model-agnostic meta-learning [62] can lead to highly accurate results in MR prostate images. However, meta-learning optimizes to obtain a convergence of the *training* process within a few steps - only indirectly influencing the number of required data points. In general, the number of epochs needed to obtain convergence of the training process is not of particular importance in biomedical imaging if it is not excessively big. In this work, we also showed that even in cases where transfer learning does not bring a big performance gain, it can still speed up training significantly (see fig. 9). Therefore, the actual benefit of meta-learning approaches compared to our approach remain unclear.

Second, multi-task learning algorithms are machine learning methods that can perform several tasks at the same time. These were also successfully applied to brain and tumor segmentation [63, 64]. For our purposes, this could mean pre-training a SFCN on age and sex prediction at the same time. It would then be interesting to re-plot the features as in fig. 1 and see whether features are now less task-specific. In this case, a transfer across tasks, even to disease prediction, might indeed be possible.

## 5 Conclusion

In this work, we proposed a transfer learning method based on the re-use of deep neural network features to alleviate the problem of domain shift across different MRI scanner sites and data sets. We showed that the performance gain can be attributed to the re-use of features across data sets. Our method achieved state-of-the-art results on age and sex prediction and led to more efficient training. Future work could combine the method with other transfer learning approaches to enable transfer to novel tasks such as disease prediction. In sum, transfer learning via re-use of deep neural network features might therefore provide a simple and efficient way of achieving highly accurate results in neuroimaging as necessary to apply deep learning in the clinic.

## Data Availability

All data produced are available online at
- http://www.ukbiobank.ac.uk
- https://fcon_1000.projects.nitrc.org/indi/abide/
- https://brain-development.org/ixi-dataset/
- https://www.oasis-brains.org/

https://fcon_1000.projects.nitrc.org/indi/abide/

https://brain-development.org/ixi-dataset/

https://www.oasis-brains.org/

## Footnotes

1 A fair comparison for age prediction would be difficult. As outlined in algorithm 1, the transfer learning process has two steps in this case (fine-tuning classifier+full model), while training from scratch has only one.

